# Preliminary epidemiological analysis on children and adolescents with novel coronavirus disease 2019 outside Hubei Province, China: an observational study utilizing crowdsourced data

**DOI:** 10.1101/2020.03.01.20029884

**Authors:** Brandon Michael Henry, Maria Helena Santos de Oliveira

**Affiliations:** Cardiac Intensive Care Unit, The Heart Institute, Cincinnati Children’s Hospital Medical Center, Ohio, USA; Department of Statistics, Federal University of Parana, Curitiba, Brazil; Pediatric COVID-19 Open Data Analysis Group, Ohio, USA

**Author notes:** **Corresponding Author:** Brandon Michael Henry, M.D., Cardiac Intensive Care Unit, The Heart Institute, Cincinnati Children’s Hospital Medical Center, 3333 Burnet Avenue, Cincinnati, Ohio, 45229, USA.

## Abstract

**Background:** The outbreak of coronavirus disease 2019 (COVID-19) continues to expand across the world. Though both the number of cases and mortality rate in children and adolescents is reported to be low in comparison to adults, limited data has been reported on the outbreak with respect to pediatric patients. To elucidate information, we utilized crowdsourced data to perform a preliminary epidemiologic analysis of pediatric patients with COVID-19

**Methods:** In this observational study, data was collected from two open-access, line list crowdsourced online databases. Pediatric cases of COVID-19 were defined as patients ≤19 years of age with a laboratory confirmed diagnosis. The primary outcomes were case counts and cumulative case counts. Secondary outcomes included days between symptoms onset and first medical care and days between first medical care and reporting. Tertiary outcomes were rate of travel to Wuhan, rate of infected family members and rates of symptoms.

**Results:** A total of 82 patients were included. The median age was 10 [IQR: 5-15] years. Patients from mainland China (outside Hubei) accounted for 46.3% of cases, while the remaining 53.7% of cases were international. Males and females accounted for 52.4% and 32.9% of cases, respectively, with the remaining 14.6% being designated as unknown. A male skew persisted across subgroup analyses by age group (p=1.0) and location (inside/outside China) (p=0.22). While the number of reported international cases has been steadily increasing over the study period, the number of reported cases in China rapidly decreased from the start point. The median reporting delay was 3 [IQR: 2-4.8] days. The median delay between symptom onset and first seeking medical care was 1 [IQR: 0-3.25] day. In international cases, time to first seeking medical care was a median of 2.5 days longer than in China (p=0.04). When clinical features were reported, fever was the most common presentation (68.0%), followed by cough (36.0%).

**Conclusions:** The number of reported international pediatric COVID-19 cases is rapidly increasing. COVID-19 infections are, to-date, more common in males than females in both the children and adolescent age groups. Additionally, this male predominance remains the case both inside and outside of China. Crowdsourced data enabled early analysis of epidemiologic variables in pediatric patients with COVID-19. Further data sharing is required to enable analyses that are required to understand the course of this infection in children.

## INTRODUCTION

The coronavirus disease 2019 (COVID-19) outbreak is rapidly expanding across the world and presents a significant public health emergency with the potential of becoming pandemic.^1^ As of March 1, 2020, cases of COVID-19 have been reported in over 60 countries. In early stages of epidemics with emerging pathogens, real-time analysis of accurate and robust epidemiological and clinical data is essential to developing interventional strategies and guiding public health decision-making.^2,3^ Such data can provide insight into infectivity, routes of transmission, disease severity, and outcomes, thus enabling epidemiologic modelling and improved public health responses.^4,5^ Line list data, which includes individual patient-level data on important epidemiological and clinical variables, is rarely available early during an outbreak with a novel pathogen.^5^ However, recognizing the utility of such data and its ability to provide for real-time analyses, multiple academic groups have been curating line list data using crowdsourcing and openly sharing the data with the scientific community.^4,5^ Crowdsourcing enables the collection of data from multiple platforms including health-care-oriented social networks, government and public health agencies, and global news sources. Sun et al. recently reported the first epidemiological analysis of crowdsourced data in 507 COVID-19 patients.^5^ However, the number of reported pediatric cases, at least in the early stages of this outbreak, is limited. In a recent analysis, of the 44,672 confirmed COVID-19 cases in China, only 2.1% were in pediatric patients (≤19 years of age).^6^ The reason for this remains unknown but may be due to underreporting of pediatric cases, as well as a combination of biologic and epidemiologic factors. During both the Severe Acute Respiratory Syndrome (SARS) and Middle Eastern Respiratory Syndrome (MERS) outbreaks, a limited number of cases with milder symptoms, fewer hospitalizations, and less mortality were observed in children as compared to adults.^7–9^

As children and adolescents represent a unique patient group, pediatric specific analyses of epidemiologic data may enable a better understanding of COVID-19 in these patients and provide situational-awareness to the pediatric health community. In this study, we analyzed pediatric cases of COVID-19 outside Hubei province, China collected from two crowdsourced curated individual line list data. We present early data on case counts, travel history, time between symptom onset, seeking medical care, and reporting, and symptomatology.

## METHODS

### Study Design

In this observational study, we collected data on children and adolescent cases of laboratory confirmed COVID-19 from two open access crowdsourced line list databases sets through March 1, 2020. Due to the format of reporting age, as well as significant lack of any age data for most patients, cases from within Hubei province of China were not able to be analyzed. As such, only cases from outside the Hubei province in China and international cases were included. However, patients who lived in Hubei and were repatriated were included in the analysis.

The methods of data curation for each of the used line lists has been previously published.^4,5^ In brief, the data curated by Sun et al. for China was collected from DXY.cn, a social network for Chinese health-care professionals, which provides real-time coverage of COVID-19 with de-identified patient data reported directly by public health agencies or state media.^5^ Each report is linked to an original source to allow for authentication. In addition, for international cases, data was curated from media sources and national health agencies. Their data is updated regularly and is freely available on the website of the Laboratory for the Modeling of Biological + Socio-technical systems website of Northeastern University. The second data set by Xu et al. (Open COVID-19 Data Curation Group) of confirmed COVID-19 cases was curated from multiple sources including World Health Organization (WHO) reports, national departments of health, and Chinese local, provincial, and national health authorities.^4^ Their data is freely available online via Google Drive.

As only de-identified patient data from publicly available databases was used in this study, no patient consent was obtained, and no ethical approval was required. This study is reported in compliance with Strengthening the Reporting of Observational Studies in Epidemiology (STROBE) Statement guidelines for observational studies (Supplement 1).

### Data Collection

Data on pediatric patients were collected from both of the line lists. This included age or age range, sex, location, date of symptom onset, date of seeking medical care, and date of COVID-19 reporting. When available, travel history, information on infected relatives, and symptomatology were also collected.

After collecting cases from both databases, overlap of patients was assessed using age, location, and the three above mentioned dates. In the event of suspected duplicate patients, one case was removed from the dataset.

### Study Definitions and Outcomes

Pediatric cases of COVID-19 were defined as patients ≤19 years of age with a laboratory confirmed diagnosis of SARS coronavirus 2 (SARS-CoV-2) infection.^5^ The definition of a laboratory confirmed cases is a patient blood or respiratory tract sample positive for SARS-CoV-2 by quantitative RT-PCR, or presence of viral genome highly homologous to reference sequence for SARS-CoV-2. Due to the format of data reporting from some Chinese provinces employing age ranges instead of raw age, the following classifications were employed for this study: children (0-≤12 years of age) and adolescents (13-≤19 years of age).

The primary outcome of this epidemiologic analysis was case counts and cumulative case counts. Secondary outcomes included days between symptoms onset and first visit to hospital or clinic, and days between first visit to hospital or clinic and reporting. Tertiary outcomes included rate of travel to Wuhan, China, rate of family members with a positive COVID-19 diagnosis, and rates of symptoms.

### Statistical Analysis

Categorical variables were reported as frequency (%) and continuous variables as median (interquartile range). Subgroups analyzed included age range, sex, and location. Case counts were compared between subgroups using Fisher’s exact test. Weekly case counts were analyzed using the date of reporting. Differences in delays between symptoms onset and first visit to hospital or clinic and days between first visit to hospital or clinic and diagnostic confirmation were compared between subgroups using Wilcoxon test. All statistical analyses were performed using R (version 3.6.1 (2019-07-05)) with a p-value of <0.05 being considered as significant.

## RESULTS

After combing the two line lists, a total of 100 pediatric cases were identified. Pediatric cases represented 3.0% of Sun et al.^5^ line list and 0.5% of the Xu et al.^4^ line list. Following removal of 18 duplicates, a total of 82 pediatric patients were included in this analysis. The date range of cases was January 21, 2020 to February 28, 2020. Patients from mainland China (outside Hubei province) accounted for 38 (46.3%) cases, while the remaining 44 cases (53.7%) were international. Within China, Yunnan province accounts for the most cases (n=8, 21.1% of cases in China). A breakdown of cases within China by province is presented in Supplement 2. Outside China, Japan has the most pediatric cases included in this analysis (n=15), accounting for 34.1% of all international cases. In total, cases from 14 countries were included (Table 1).

**Table 1.** Geographical Distribution of Pediatric COVID-19 Cases (n=82).

| Country | Pediatric Cases of COVID-19 |  |  |
| --- | --- | --- | --- |
|  | Age 0-12 | Age 13-19 | TOTAL |
| Australia | 1<br>(100.0%) | - | 1 |
| China | 25<br>(65.8%) | 13<br>(34.2%) | 38 |
| France | 1<br>(100.0%) | - | 1 |
| Hong Kong | - | 1<br>(100.0%) | 1 |
| Italy | 3<br>(42.9%) | 4<br>(57.1%) | 7 |
| Japan | 10<br>(66.6%) | 5<br>(33.3%) | 15 |
| Malaysia | 4<br>(100.0%) | - | 4 |
| Singapore | 4<br>(80.0%) | 1<br>(20.0%) | 5 |
| South Korea | 1<br>(50.0%) | 1<br>(50.0%) | 2 |
| Spain | - | 1<br>(100.0%) | 1 |
| Switzerland | ** | ** | 2 |
| Thailand | 2<br>(100.0%) | - | 2 |
| United Arab Emirates | 1<br>(100.0%) | - | 1 |
| Vietnam | 1<br>(50.0%) | 1<br>(50.0%) | 2 |
| **TOTAL** | **53 (%)** | **27 (3%)** | **82** |

\*Data presented as n= (%).
\*\*No age data was available on the cases from Switzerland.

The median age of patients was 10 [IQR: 5-15] years. The distribution of patients by age is presented in Figure 1. No significant differences in age were detected between cases inside and outside China (10 [IQR: 5-15] inside vs. 9 [IQR: 5-15.5] outside; p=0.91). For 13 patients (15.9%), only an age range was available. To include these patients in further analyses, cases were split into two age groups based on how data was reported in the line lists. There were 53 children (65.6%) between 0-12 years and 27 adolescents (32.9%) between 13-19 years, with 2 missing age range data (2.4%).

**Figure 1.**
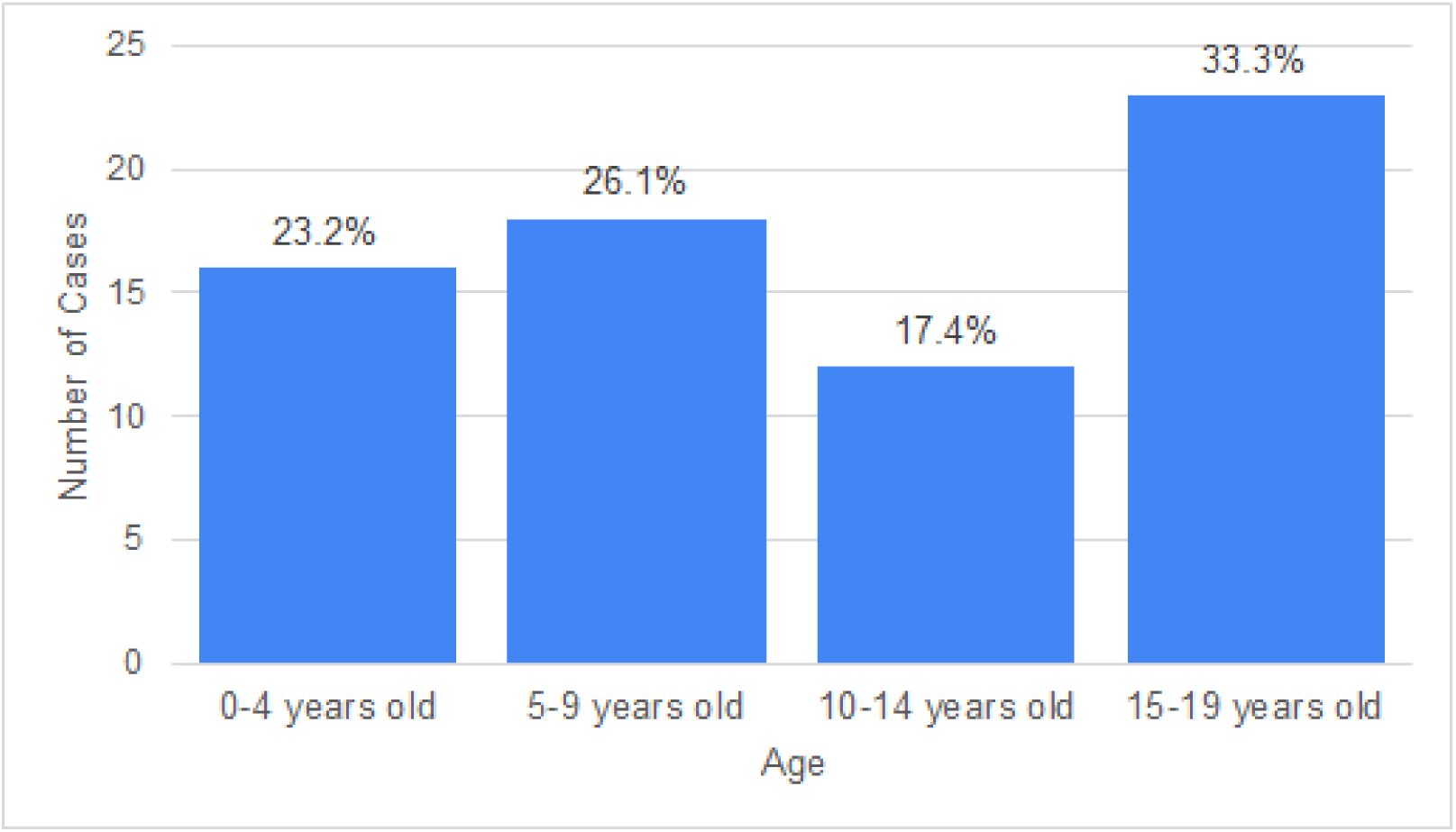
Age distribution of COVID-19 patients (n=69).

Males accounted for 52.4% of included cases, females accounted for 32.9% and 14.6% were unknown (Table 2). When broken down by age range, no significant differences were observed between the proportion of cases between age groups (p=1.0), with a male skew of cases seen within both (Table 2). When compared by location, males accounted for 52.6% of cases within China and 57.3% outside China (p=0.22).

**Table 2.**
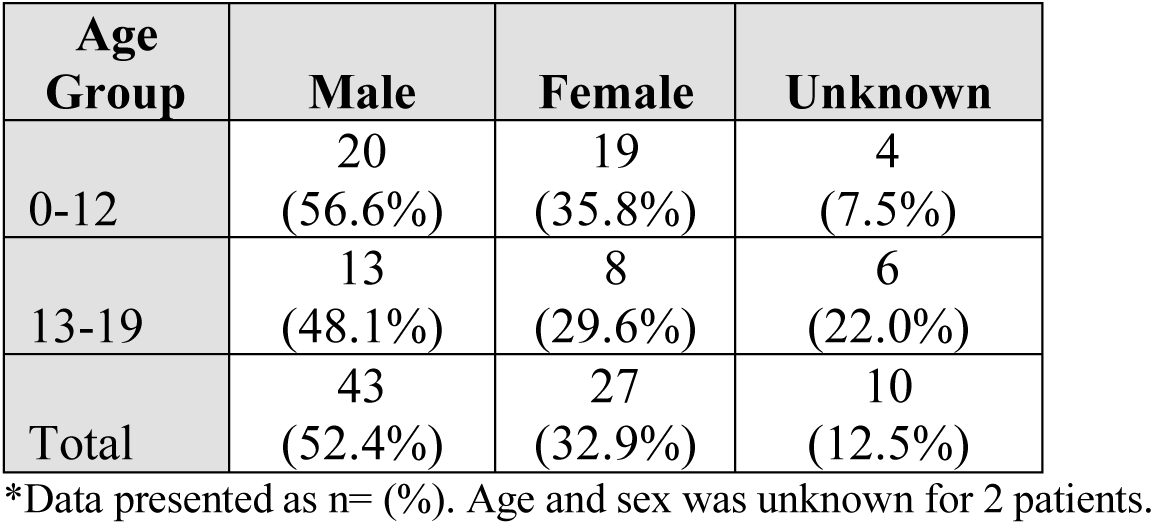
Sex Distribution of Pediatric COVID-19 Cases.

| **Age Group** | **Male** | **Female** | **Unknown** |
| --- | --- | --- | --- |
| 0-12 | 20<br>(56.6%) | 19<br>(35.8%) | 4<br>(7.5%) |
| 13-19 | 13<br>(48.1%) | 8<br>(29.6%) | 6<br>(22.0%) |
| Total | 43<br>(52.4%) | 27<br>(32.9%) | 10<br>(12.5%) |

\*Data presented as n= (%). Age and sex was unknown for 2 patients.

A timeline of cases and cumulative cases is presented in Figure 2 by age range and Figure 3 by location. In both children and adolescents, weekly cases peaked between January 20-26, 2020. The number of reported cases continued to drop reaching a low in mid-February, then undergoing a rapid increase sustained through to the end of the study period, driven by new international cases. Within China, the number of weekly cases steadily decreased from the beginning of the study, with no new cases added to the databases since February 3-9, 2020. The number of cumulative international cases has been constantly increasing over the study period with a more rapid increase in recent weeks.

**Figure 2.**
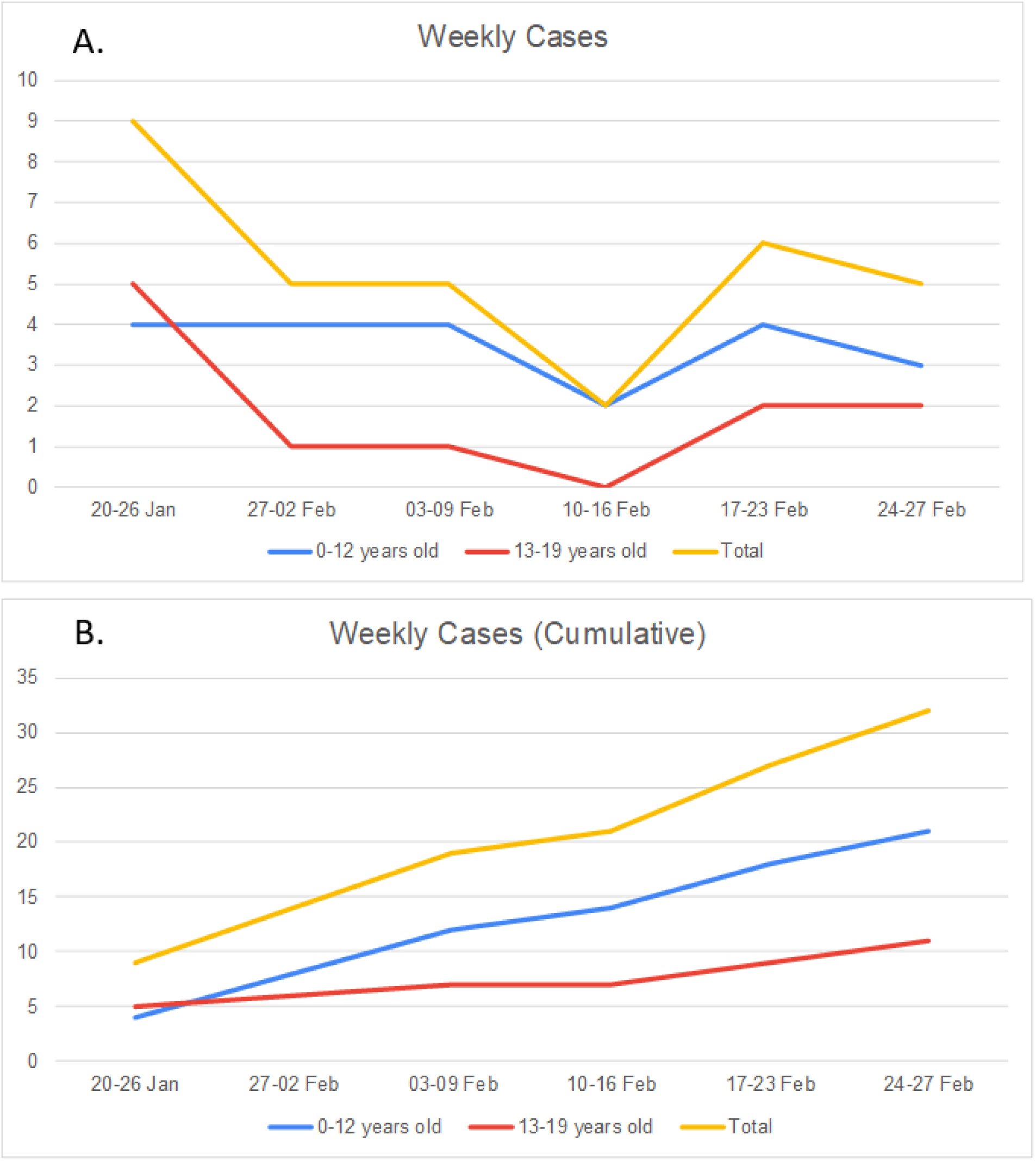
Number of Weekly Cases (A) and Cumulative (B) Weekly Cases of Pediatric COVID- by Age Group.

**Figure 3.**
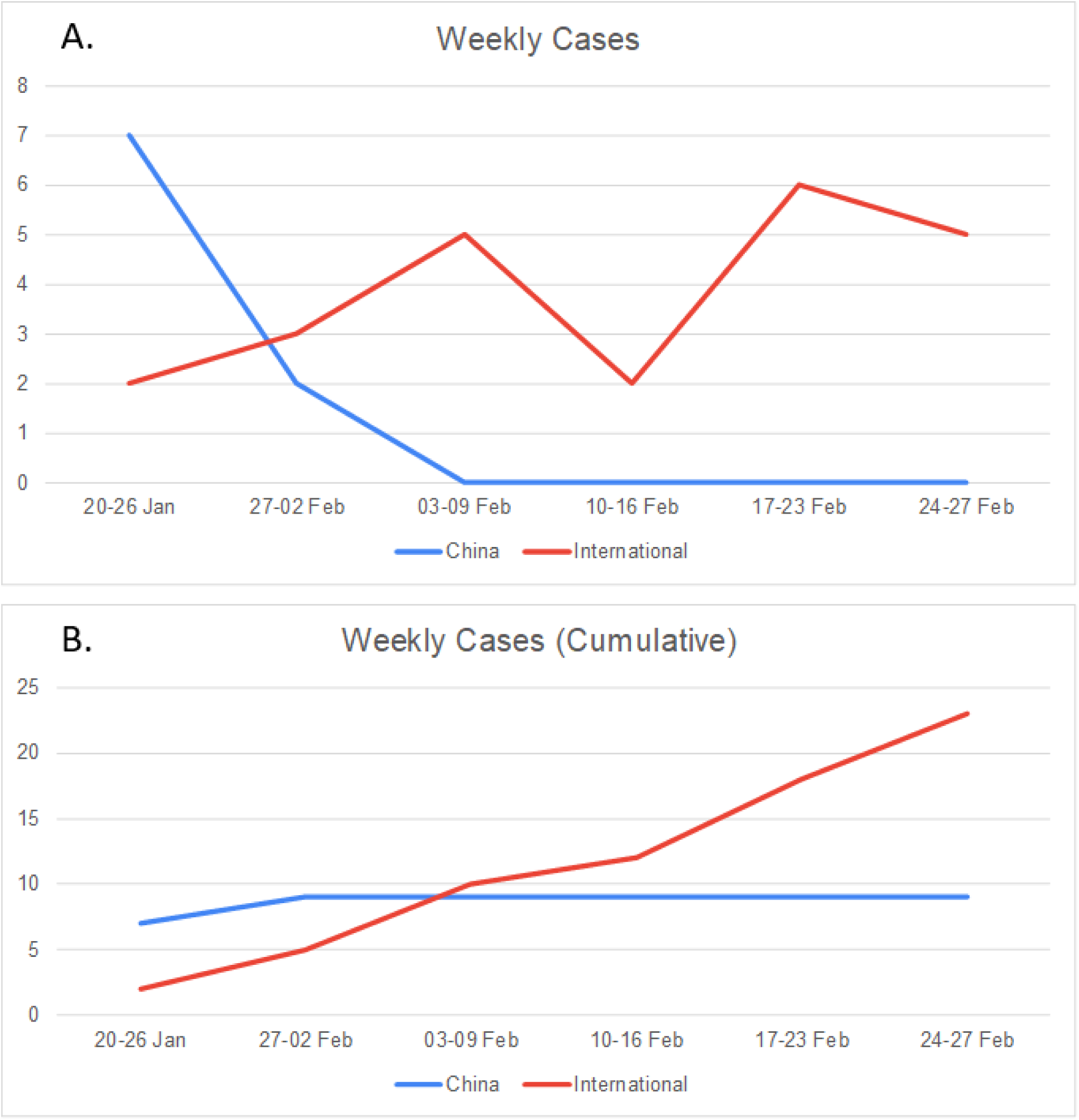
Number of Weekly Cases (A) and Cumulative (B) Weekly Cases of Pediatric COVID- by location inside versus outside China.

The median delay between first visit to hospital/clinic and reporting was 3 [IQR: 2.0-4.8] days (n=14). No significant differences in this delay time was observed for subgroups of age (p=0.29) or location (inside/outside China) (p=0.67).

The median delay between the onset of symptoms and first visit to hospital/clinic was 1 [IQR: 0-3.3] day (n=20). There was no significant difference with respect to age subgroup (p=0.93). However, within China this delay was significantly shorter with a median duration of 1 day [IQR: 0-2.5] versus 3.5 [IQR: 1.5-7] days abroad (p=0.04).

A total of 14 (17.1%) cases were in patients who lived in Wuhan, China and were repatriated. Among the remaining 68 cases, 25 (30.5%) patients had a history of travel to Wuhan. No information on travel history was available for the other 43 (52.4%) patients.

A total of 29 (35.4%) patients were noted to have an infected family member. In one adolescent case in Japan, the patient was the son of a physician who had earlier tested positive for the virus. Data on patient symptomatology was available in 25 (30.5%) of the cases. Fever was the most common symptom, present in 17 (68.0%) of those cases, while 2 (8.0%) cases were asymptomatic. Cough was reported in 9 (36.0%) cases. A breakdown of patient symptoms is presented in Table 3. Data on disease severity, need for hospitalization or intensive care support, and patient outcome was not available for analysis.

**Table 3.** Reported Symptoms in Pediatric COVID-19 Cases (n=25).

| Symptom | Number of Cases |
| --- | --- |
| Fever | 17 (68.0%) |
| Cough | 9 (36.0%) |
| Pharyngitis | 3 (12.0%) |
| Rhinorrhea | 2 (8.0%) |
| Abdominal Pain | 1 (4.0%) |
| Diarrhea | 1 (4.0%) |
| Malaise | 1 (4.0%) |
| Asymptomatic | 2 (8.0%) |

\*Data presented as n= (%).

## DISCUSSION

Real-time analysis of crowdsourced individual line list data provides a tool for monitoring outbreaks with novel pathogens in early stages, as witnessed with Ebola.^5,10^ Though this data is often difficult to obtain at the start of an outbreak, thanks to the efforts of multiple academic teams in curating line list data from multiple sources, we have been able to gain some insight into the current COVID-19 outbreak in children.^4,5^ Though this data cannot replace official data and analyses provided by national and international health agencies, it can enable rapid analysis of data and dissemination of key information in the preliminary stages of an outbreak when minimal other data is available, and, as applied in this study, early data on specific subsets of patients.^5^

This study combined data from two crowdsourced line lists to obtain a sample of pediatric patients with COVID-19 in order to gain some preliminary insights into the outbreak in children and adolescents. Cases in these patients represented only a small proportion of each line list.

Reports from both the Chinese Center for Disease Control and Prevention and the WHO on the current COVID-19 outbreak, have found pediatric patients represent a small proportion of all cases, with an attack rate of 2.4%.^6,11^ Children with COVID-19 appear to have milder symptoms as noted by a severe case rate of 2.5%, with only 0.2% developing critical illness.^11^ Similar finding of limited cases and milder symptoms in children was observed in both SARS and MERS outbreaks.^7–9^ However, whether the limited number of pediatric COVID-19 cases is due to less susceptibility or milder presentation leading to missed detection remains unknown. Of note, initial screening criteria of COVID-19 in China focused on cases of suspected pneumonia.^12^

The mild nature of pediatric cases is likely multifactorial. Age related differences in innate immunity^13,14^, in which monocytes and macrophages may play an important role controlling an emerging viral pathogen, may be partially responsible for the milder observed symptoms and asymptomatic cases. Others have suggested that respiratory tract health, including less exposure to pollution, as well as lung epithelial cell senescence and immunosenescence of immune cells of the respiratory system may play a role.^15^ Cai et al. reported that while COVID-19 viral shredding in respiratory specimens is longer in children than adults and also observed in the stool specimens, the virus was not detected in serum samples.^16^ The absence of viremia likely contributes to the lack of serve illness in pediatric cases.

In both MERS and SARS outbreaks, children were less likely to develop severe respiratory distress seen in adults, experiencing a pneumonia or common cold symptoms.^7–9^ Exposure of young children in China to other coronaviruses, such as OC43, 229E, NL63, and HKU1,^17^ may impart some immunity not reinforced in older individuals, explaining partial protection against COVID-19 in this population.^18,19^

Comorbidities have been reported to be associated with severity of COVID-19 infection.^20,21^ As children generally have less comorbidities, this may also in part explain the lack of serious infections in children and contribute to decreased detection. Moreover, behavioral differences and limited travel as compared to adults may have also explained the limited number of cases early in the outbreak.^5^

In our data set, children under 10 years old made up a near equal proportion of included cases. This is different than data presented in a recent Chinese CDC report which showed a slightly greater number of cases in those over age 10 (416 aged 0-9 vs. 549 aged 10-19).^6^ Whether this is due to the availability of cases in the line lists or a higher detection of cases in young children outside China is unclear, however, this should be monitored going forward. In general, infants and young children are often at increased risk of poor outcomes from respiratory infections,^22^ which has not yet been witnessed with COVID-19 from reported aggregate data.^6^

Interestingly, a significant male skew in cases was observed in pediatric patients throughout the analysis. This sex skew was not observed among children in SARS or MERS. In adults, cases of COVID-19 are reported to be skewed towards men also.^5,6^ However, explanations applied to adults, such as the high prevalence rates of smoking^23^ and comorbidities (i.e. cardiovascular disease and diabetes)^24,25^ in Chinese men as compared to women, would not apply to children. Though adult females produce more robust inflammatory responses as compared to men, the opposite is true in infants and children.^26^ Young males produce stronger inflammatory responses than females.^26^ However, females, beginning at birth and persisting throughout adulthood, have higher numbers of CD4+ T cells and higher CD4/CD8 T cell ratios^26^, though the potential impact of this on COVID-19 infections is unknown.

In respiratory syncytial virus (RSV) related illness, a common respiratory tract infection in young children, prevalence rates are reported to be the same in males and females, however, males are more likely to suffer from more severe infections requiring hospitalization.^27–29^ We hypothesize that a similar pattern may be observed with COVID-19, with males developing more symptoms, thus leading to increased detection and reporting in these patients.

In this analysis, the male skew persisted through subgroup analysis by age, with no difference in the rate of infected males in children versus adolescents. As such, the impact of the protective effects of estrogen receptor signaling in females observed in SARS may be minimal,^30^ as we expect a more balanced sex ratio in younger children as compared to older pubescent females.

To account for the skew in the sex ratio in the Chinese population as a potential confounder, we compared the rate of cases by sex inside and outside China, finding no significant differences. Larger epidemiologic studies are needed to confirm this sex skew in children and adolescents COVID-19 cases and, if persistent, further investigations are needed to understand this phenomenon.

The weekly counts of reported cases in this study showed unique trends based on geographical region. In China, a rapid drop from a peak at study initiation on January 20 to no new cases after February 9 was observed. Within Wuhan, the epicenter of the outbreak, all notifiable cases after January 22 were in patients ≥ 15 years of age.^20,21^ In the study by Sun et al., they used January 18 as the date when media and public awareness of COVID-19 became more pronounced, followed by significant governmental efforts aimed at containment.^5^ No cases in this data set were reported before this date. Whether the decrease in new cases in China is due to reporting difficulties/lack of case inclusion in the line lists or effectiveness of Chinese public health interventions aimed at slowing the spread of the virus, or a combination of both, is unclear.

Internationally, the number of reported cases has steadily increased with a faster pace in recent weeks. We expect that this trend will continue, especially as testing becomes more readily available in new countries. In Figure 3A, a small dip in reported cases in the last few days of February is observed. We believe that this is primarily due to the delay in reporting, which was calculated to be a median of 3 days from the first medical visit in this study. As such, we expect to see the number of cases in the last week increase in the coming days.

Though analyzed based on a small subset of included patients, patients in China were found to first receive medical attention a median of 2.5 days sooner after initiation of symptoms than those abroad. We hypothesize that this is due to heightened awareness of COVID-19 in China and perception of decreased risk of COVID-19 infection among those abroad. However, this is opposite of the trend observed by Sun et al., which may be due to a higher number of international travelers in their data set.^5^

Data on patient symptoms was only available for 25 patients. Fever was the most common presentation, followed by cough, with only 2 being asymptomatic. However, how representative this small sample is remains unclear. Moreover, as this data was not collected directly for the purpose of this analysis, we cannot verify if a systematic review of symptoms was performed. Nonetheless, our data was similar to a case series of COVID-19 in 10 children, which observed fever in 80% and cough in 60%. While the WHO has noted that COVID-19 appears to present differently from SARS, fever followed by cough, was the most common presentation of SARS in children.^7,11^ Further progression of COVID-19 illness in children developing more severe symptoms requires further data and analysis from the limited number of severe cases reported.

The WHO Joint Mission reported that many pediatric cases were identified through contact tracing in households with infected adults and reported many pediatric patients had only mild symptoms.^11^ A report by Chan et. al on COVID-19 clusters found in one household that 5 adults had symptomatic infections, while a child present in the residence tested positive for COVID-19 but was asymptomatic.^31^ An infected family member was commonly reported in the present study, including a case of a physician transmitting the infection to his son. In another report, household exposure was reported found to be 70% in 10 pediatric COVID-19 patients.^16^

Based on the obtained data, we suggest that in the presence of active community transmission in a region, children presenting with a fever of unknown cause or with a fever in the presence of common cold or pneumonia symptoms should be tested for COVID-19 and influenza A and B. Additionally, as transmission between child to adult has been reported^16^, pediatric healthcare providers should take appropriate precautions when caring for a child with suspected COVID-19. This study has several limitations. First, it presents data from a limited sample of total pediatric cases of COVID-19. As noted by Sun et al., line list data is available for only a small number of Chinese provinces, while aggregated data on number of cases and mortality is provided by all.^5^ As such, cases from some provinces, including Hubei which is the center of the initial outbreak, were not able to be included. Cases in this study may also be those in whom the disease was severe enough to seek medical attention or identified by contact tracing, and as such, many minor cases may have been missed and symptomatology data skewed.^5^ Additionally, crowdsourced data would be less likely to capture mild cases as curated via the applied data collection modalities.^5^ Missing data was common for many of the included variables. Further efforts should be made to collect follow up data on patient outcomes.

Limited data in children with respect to travel history or exposure prevented any estimation of incubation time. Sun et al. reported a median incubation time of 4.5 days [IQR: 3.0.5.5]^5^, while others using their data and other data sets have reported incubation rates of 5-6 days.^21,32–34^ In a case series of 10 children, Cai et al. observed a longer incubation than observed in adults, with a mean of 6.5 days (range 2-10 days) from exposure to symptoms.^16^ Further efforts should be made in the coming weeks to study transmission and estimate incubation time in pediatric patients to improve public health guidance.

As the outbreak progresses and the curated line lists continue to grow, further analyses of pediatric epidemiologic data should be performed regularly. To that end, the authors have established the Pediatric CO-VID19 Open Data Analysis Group, an international collaborative effort to study the disease in children and adolescents. However, line lists curated by academic teams will struggle to keep pace with the rapid increase in number of cases as COVID-19 trends towards pandemic. We encourage coordinated efforts between national and international health agencies and academia to produce line lists of patients which in turn will better enable the medical community to develop effective interventions against COVID-19.

## CONCLUSIONS

The number of reported COVID-19 cases in pediatric patients is rapidly increasing around the world. To date, COVID-19 infections are significantly more common in males than females in both children and adolescents inside and outside of China. When symptomatic, fever is the most common presentation, with or without symptoms of a common cold. In this study, crowdsourced data enabled early analysis of epidemiologic variables in pediatric patients with COVID-19. Analyses of larger datasets are urgently needed to better understand this infection in children and provide further situational awareness to the pediatric health community.

## Data Availability

The line lists used in this study are freely available online at the website of the Laboratory for the Modeling of Biological + Socio-technical Systems of Northeastern University and Google Drive.
Those interested in our pediatric specific dataset can email the corresponding author for data. The Pediatric CO-VID19 Open Data Analysis Group, a working group established to analyze pediatric epidemiologic and clinical data on COVID-19, strongly welcomes collaboration and participation from the international medical and scientific community. Those interested are encouraged to contact the corresponding author.

https://www.mobs-lab.org/

https://tinyurl.com/s6gsq5y

## DATA SHARING

The line lists used in this study are freely available online at the website of the Laboratory for the Modeling of Biological + Socio-technical Systems of Northeastern University and Google Drive. Those interested in our pediatric specific dataset can email the corresponding author for data. The Pediatric CO-VID19 Open Data Analysis Group, a working group established to analyze pediatric epidemiologic and clinical data on COVID-19, strongly welcomes collaboration and participation from the international medical and scientific community. Those interested are encouraged to contact the corresponding author.

## ACKNOWLEDGEMENTS

The authors would like to thank everyone involved in the curation of the line list datasets that enabled this study. The results and conclusions in this study are those of the authors and do not represent the official position of their respective institutions. The authors declare no conflicts of interest.

## FUNDING

None.

## SUPPLEMENT LEGENDS

**Supplement 1. STROBE Statement Checklist**

**Supplement 2. Distribution of Included Pediatric COVID-19 Cases in Chinese Provinces (n=38)**.

## Notes

### Competing Interest Statement

The authors have declared no competing interest.

### Funding Statement

No external funding.

